# SynTrustBench: Benchmarking Synthetic Clinical Data with Calibrated False-Positive Rates and Certified Limits of Detection

**DOI:** 10.64898/2026.08.05.26359803

**Authors:** Neeam Shahriar Hayder, Behraj Khan, Syed Ahmad Chan Bukhari

## Abstract

Medical research requires large volumes of patient data. Such data are difficult to obtain because of strict privacy regulations. Artificially generated data (synthetic data) promise to share data safely, thereby bypassing any need to go through strict, time-consuming regulations. To test this promise, benchmarks have been developed to evaluate synthetic data on their safety and usefulness. However, the benchmarks themselves are rarely evaluated. We propose **Syn-TrustBench**, an executable framework for evaluating the fidelity, utility and privacy of synthetic clinical data. Unlike existing benchmarks, SynTrustBench does not assume its metrics are valid before testing them. For each dimension it evaluates, SynTrustBench estimates two properties. Its false-positive rate is controlled by setting the pass/fail threshold at the 95th percentile of the null distribution from 100 real-versus-real comparisons, so real data of the calibration size fail at most 5% of the time. Its limit of detection is estimated by injecting failures of known type and size, such as copied training records, at graded doses into real data; it is the smallest dose detected with at least 95% probability, certified at 95% confidence. Every SynTrustBench verdict therefore carries a confidence statement. On two public diabetes datasets, real data failed in 2.8% and 3.2% of real-versus-real comparisons, within the 5% bound, and every certified limit of detection held on fresh data. SynTrustBench detected as few as 3 copied training records among 11,443, and, in an exploratory application to real generators, it flagged a privacy-relevant proximity anomaly in a TVAE generator on every seed that a standard membership-inference attack did not register (AUC 0.50). Against SDMetrics under the same calibration, it matched or exceeded the privacy metric on one dataset but not the other, and was more sensitive on fidelity.

**CCS Concepts:**

- **Applied computing** → **Health informatics**; • **Security and privacy** → *Human and societal aspects of security and privacy*; • **Computing methodologies** → *Machine learning*.

## 1 Introduction

Patient-level clinical data are hard to share: privacy, consent and institutional rules all stand in the way. Synthetic data are proposed as a way around this: artificial tables that preserve the statistical patterns of the source data while promising protection against inference attacks [5, 19]. Whether a given synthetic table can be released is decided by an evaluation. Does it resemble the data distribution it came from? Does a model trained on it work on real patients? Could a real patient be traced back? Benchmarks and toolkits answer these questions with dozens of metrics [10, 25, 29, 32, 43].

The validity of the metrics themselves is rarely tested, and three problems recur. First, *a pass can be a false negative*. A release consisting entirely of real held-out records passes similarity-based privacy metrics such as distance to closest record (DCR) and nearest-neighbour distance ratio (NNDR) [13]. In earlier work [17], a CT-GAN table reproduced a training record verbatim, yet a distance-based membership-inference attack achieved an AUC of 0.497 (95% CI 0.489–0.505), indistinguishable from chance. Second, *thresholds are not referenced to a null distribution*. A fixed cut-off ignores how much two independent samples of real patients already differ, so its false-positive rate is uncontrolled and varies across datasets. In concurrent work on the same cohort, a gate built from fixed thresholds failed or refused every real-versus-real comparison [18]. Third, *missing evidence is silently omitted*. In a focused audit of 12 structured-tabular clinical reports, all 12 evaluated fidelity, one evaluated equity, and none evaluated all five dimensions considered here (Table 1). A dimension that was not measured is absent from the result rather than reported as missing, so a reader cannot distinguish “not evaluated” from “passed”.

**Table 1:** Evidence reported in a focused audit of structured-tabular synthetic clinical data reports (*N* = 12) [17].

| Evidence component | Reports |
|---|---|
| Fidelity evaluation | 12/12 |
| Held-out-real utility | 11/12 |
| Privacy evaluation | 10/12 |
| Equity or subgroup evaluation | 1/12 |
| Robustness evaluation | 4/12 |
| Uncertainty reported | 6/12 |
| Code available | 7/12 |
| Evaluated all five dimensions | 0/12 |

A practitioner reading “privacy: pass” therefore cannot tell whether the table is safe or the metric lacked the sensitivity to detect a leak. Clinical laboratories face the same problem and address it through analytical validation. Before an assay is used, its specificity is established on samples known to be negative, and its analytical sensitivity is reported as a limit of detection: the lowest concentration detected with at least 95% probability [7]. A negative laboratory result is therefore read as “below the limit of detection”, not “absent”. We argue that synthetic-data benchmarks should be validated in the same way, so that a pass is read as “no failure above the stated limit of detection”.

*This paper*. We introduce SynTrustBench (STB) v0.5, a benchmark for synthetic clinical tables in which every verdict is tested before it is trusted. Our contributions are:

1. **Calibrated verdicts**. Five dimensions are measured; fidelity, utility and privacy decide the verdict through the null-calibrated gate of concurrent work [18], which we adopt rather than propose. Its threshold is set from honest real-versus-real comparisons and bounds the false-positive rate of the whole gate for tables of the calibration size (Proposition 1); each dimension returns Pass, Fail or Not Evaluated, so missing evidence stays visible (Section 3).
2. **Verdicts that state what they rule out**. Every Fail carries its strength of evidence, and every Pass carries a limit of detection for each kind of planted flaw: the smallest dose caught with at least 95% probability, certified with 95% confidence (Proposition 2).
3. **A test of the benchmark, and of its competitors**. On two public diabetes datasets we plant eleven families of flaws, nine of them at graded doses, into honest tables, check the stated limits on fresh data, and compare STB with SDMetrics under the same calibration rule (Sections 4 and 5).
4. **A preregistered, reproducible release**. The design, hypotheses and predictions were fixed before scoring; every number is regenerated from saved evidence by one script, released at https://anonymous.4open.science/r/SynTrustBench_v05/ (Appendix A).

Earlier work [17] introduced the five-dimension evaluation and the audit of Table 1; concurrent work [18] proposes and evaluates the calibrated gate and how evidence from several dimensions should be *combined*. New here are the limits of detection, their certification, validation on fresh data and transfer to generator output, and the matched comparison with SDMetrics.

## 2 Background and Related Work

### Evaluation frameworks

Reviews and frameworks for synthetic health data differ in scope, terminology, aggregation, tasks and attackers [19, 22, 27, 31, 39, 44]. Benchmarks such as the multifaceted EHR benchmark of Yan et al. rank generators against each other [43]; toolkits such as synthcity, SynthEval and SDMetrics compute many metrics [10, 25, 32]; NIST’s SDNist reports deidentification quality [29]. Belgodere et al. use the same five-part vocabulary for auditing [1]. These tools measure; they rarely say how much a measurement can be trusted. A ranking without an absolute floor cannot say whether the best generator is good enough, and a metric without a validated detection range cannot say what its pass excludes.

### Critiques of the metrics

Similarity-based privacy metrics, such as distance to closest record, can be passed by data that copy real records, and their outputs can themselves leak training records [13, 14]. Outliers are the records synthetic data protect least [35]. Utility conclusions depend on which downstream model is chosen, and generator rankings change once hyperparameters are tuned [12]. Fidelity and diversity metrics fail basic sanity checks [33]. Empirical privacy tests give lower bounds on risk, not guarantees: failing to find a leak is weak evidence that none exists [11]. STB takes these critiques as design constraints rather than as reasons to abandon measurement.

### Planted flaws and limits of detection

Privacy auditing plants known records (*canaries*) and checks whether they can be detected [36]. Clinical laboratories validate assays the same way: replicates of a dilution series give a hit rate at each concentration, and the limit of detection is the concentration detected with a stated probability, conventionally 95% [7, 41]. STB applies this logic to every check.

### Audit

We analyzed the structured-tabular subset of a frozen 30-report corpus: reports that generated or evaluated synthetic clinical data and reported at least one quantitative fidelity, utility or privacy assessment [2, 4, 6, 15, 16, 20, 21, 23, 24, 28, 34, 38]. Evaluation concentrated on fidelity, utility and privacy (Table 1). Privacy methods also varied: seven reports used membership inference, three attribute inference, two re-identification or reconstruction, and two documented a verified differential-privacy guarantee. The coding was done by one author and describes this corpus, not the field.

## 3 SynTrustBench v0.5

Figure 1 summarizes the protocol: thresholds are calibrated on honest real-versus-real comparisons, their sensitivity is measured with planted flaws, and real generators are then judged by the same test.

**Figure 1:**
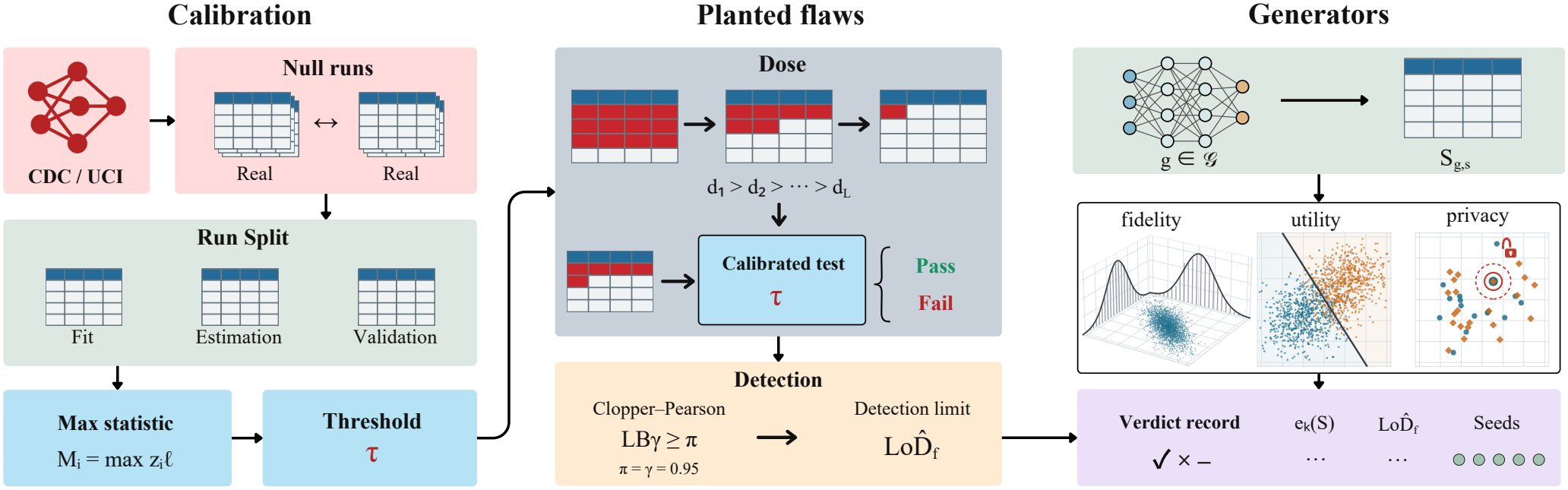
The SynTrustBench protocol. *Calibration:* null runs compare real data with real data; the threshold τ is the 5th largest maximum statistic *M*_*i*_ over the 100 fit runs, bounding the false-positive rate by 5/101 for tables of the calibration size. *Planted flaws:* flaws at decreasing doses give the limit of detection 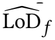, certified at π = γ = 0.95. *Generators:* each synthetic table is judged by the same test and receives a verdict recorsd.

**Figure 2:**
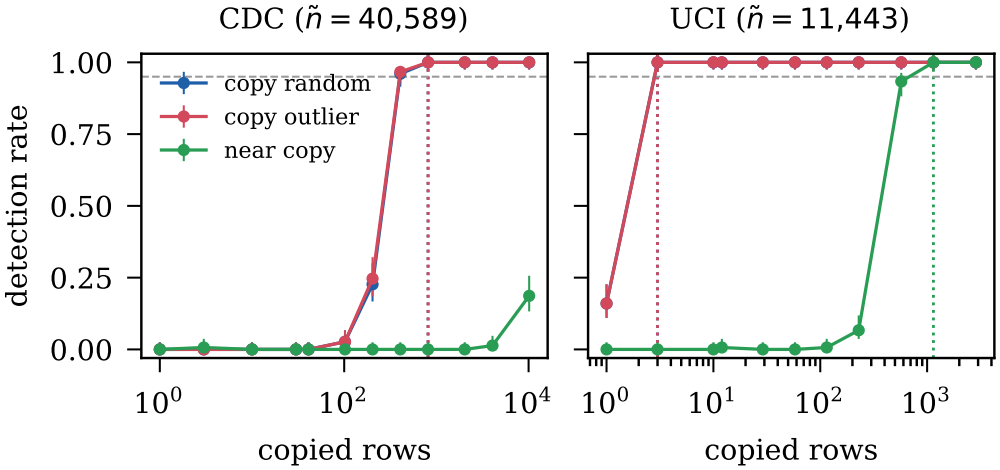
Detection rate of STB’s gate against the number of planted rows, for the privacy families on honest tables (estimation bases; 150 replicates per dose). Bars: Wilson 95% intervals. Dashed line: 0.95. Dotted lines: certified limits of detection.

### 3.1 Data roles and the null hypothesis

Let *D* be a real cohort with one row per person. STB splits *D* once, stratified by outcome and disjoint by hashed person identifier, into a generator-training set *T* (40%), a calibration pool *P* (40%) and a held-out test set *E* (20%). Because each row is one person (the earliest encounter in UCI; one respondent in CDC), row-level resampling is person-level resampling. The pool is halved; one half is the fidelity reference *R*, with *n* = |*R*| rows. A synthetic table *S* is generated from *T* with |*S*| = *n* rows and is judged against three real roles: *T* (the privacy member population), *R* (the fidelity reference) and *E* (the utility test set).

For each table, STB tests the null hypothesis *H*_0_ that *S* is distributed as an independent sample of *n* real patients from the population of *T* . A rejection therefore means *a departure from real data that is detectable at this sample size*, not that the table is harmful.

#### Null runs

To approximate the distribution of every statistic under *H*_0_, STB draws *null runs*. Run *j* re-splits the development data *T* ∪ *P* with a fresh seed, in the same 40/40/20 proportions, into a pseudo-training set 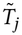, a pseudo-pool that is halved into a reference 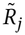 and an honest table 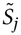 of equal size 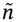, and a pseudo-test set 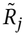; all four are person-disjoint. Every statistic below is computed for *S*_*j*_ exactly as for a synthetic *S*, and *E* is never read. Because the development data are 80% of *D*, null roles are 80% the size of the main roles (*ñ* = 0.8*n*). Planted-flaw experiments use null runs as their bases, so their candidates match the null runs in size exactly; real generator tables are judged on the main roles and are 1.25 times larger. Proposition 1 therefore applies exactly to tables of the null-run size; for generator tables the bound is an approximation, which this study does not test at full size (Section 7).

### 3.2 Signals

Each gating dimension *k* ∈ {fid, util, priv} is measured by a vector of signals *x*_*k*_ (*S*), each oriented so that larger values indicate a larger departure from *H*_0_. Where a statistic has a non-zero real-data value, the signal is the candidate’s value minus the *floor*: the same statistic computed with a real table in the candidate’s place. Full definitions are in Appendix C.

#### Fidelity (reference R)

(i) *Classifier two-sample test:* a gradient-boosted classifier is trained to separate *R* (label 0) from *S* (label 1) with 5-fold cross-fitting, and AUC_det_(*S*) is computed on out-of-fold scores; the signal is AUC_det_(*S*)−AUC_det_(*T* ). (ii) *Coverage deficit:* for categorical column *c*, cov_*c*_ (*S*) = |*R*|^−1^ ∑_*r*∈*R*_ **1**[*r*_*c*_ ∈ levels_*c*_ (*S*)], the share of reference patients whose category appears in *S*; the signal is max_*c*_[cov_*c*_(*T* ) − cov_*c*_ (*S*)]. (iii) *Marginal distances:* the mean over continuous columns of 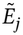, and the mean base-2 Jensen–Shannon divergence over categorical columns, each minus its floor. (iv) *Association error:* the mean absolute difference between the pairwise association matrices of *R* and *S* (absolute Pearson correlation, Cramér’s *V*, or the correlation ratio, by variable type), minus its floor. (v) *Clinical-rule violations:* the largest per-rule violation rate in *S*; real data violate no rule.

#### Utility (test set E)

Three probes *h* ∈ {random forest, logistic regression, gradient boosting} are trained on *S* and on *T* with pre-processing fitted on *T*, and evaluated on *E*. The chance-corrected AUC retention is

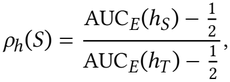

which is 1 when the synthetic-trained probe matches the real-trained one and 0 at chance, whatever the real-data AUC. It is computed only when the lower 95% bootstrap bound of AUC_*E*_(*h*_*T*_ ) exceeds 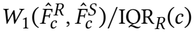; otherwise utility is Not Evaluated. The signal is the *upper* limit of a 95% paired percentile bootstrap interval over rows of *E* (*B* = 200), oriented so that low retention is harmful; a table is penalized only to the extent that even its most favourable plausible retention is low. Probability calibration of the logistic probe enters through the ratios of calibration-in-the-large 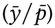 and calibration slope for *h*_*S*_ relative to *h*_*T*_ ; each ratio contributes two oriented signals, so departures in either direction count. Within every role assignment the candidate and the reference have equal size, and null runs use the same construction, so the difference in training size between *h*_*S*_ and *h*_*T*_ is represented in the null distribution (up to the scaling noted in Section 3.1).

#### Privacy (members T, non-members E)

Rows are encoded with standardized continuous columns and one-hot categories fitted on *T*, and compared by Euclidean distance. (i) *Exact-match excess:* 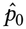, where 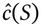 is the fraction of rows of *S* identical to some row of *T* (all rows used) and *p*_0_ is the fraction of rows of *E* identical to some row of *T*, the rate at which distinct real people coincide by chance. (ii) *Proximity share:* the fraction of rows of *S* whose nearest-neighbour distance to *T* is below the 5th percentile of non-self nearest-neighbour distances within *T* . (iii) *Membership-inference AUC:* the AUC of the score *s*(*x*) = − min_*z*∈*S*_ ‖ϕ(*x*) − ϕ(*z*)‖_2_ for separating members (*x* ∈ *T* ) from non-members (*x* ∈ *E*). Nearest-neighbour statistics use reproducible samples of at most 10,000 rows per role; the exact-match count uses every row. All three are empirical, attacker-specific indicators: failing them flags a privacy-relevant anomaly that warrants investigation, not demonstrated leakage, and passing them is not a privacy guarantee [11, 13].

#### Diagnostic dimensions

Equity records the largest representation gap of a protected attribute between *R* and *S* and the worst sub-group retention min_*g*_ ρ^(*g*)^ over subgroups with at least 30 test patients and at least 10 of each outcome. Because the minimum of several noisy estimates is biased downward and grows more extreme with the number of groups, equity is calibrated like every other dimension (its null runs contain the same number of groups) but does not decide the verdict unless admitted (Section 3.5). Robustness, the stability of utility under resampling, added missingness and reduced size, is reported likewise.

### 3.3 Calibrated verdicts

Let *x*(*S*) ∈ ℝ^*p*^ concatenate the gating signals and let *x*_1_, …, *x*_*m*_ be those of the first *m* = 100 null runs (the *fit* runs). Pool the candidate with the fit runs, *x*_*m*+1_ := *x*(*S*), and standardize each coordinate *l* by the pooled median and standard deviation,

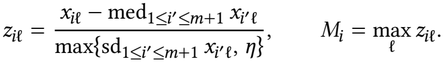

With *r* = ⌊α(*m* + 1)⌋, let τ be the *r* -th largest of *M*_1_, …, *M*_*m*_. Dimension *k fails* if max_*l*∈*k*_ *z*_*m*+1,*l*_ > τ, passes otherwise, and is *Not Evaluated* if any of its required signals is unavailable (for example, utility when the real-trained probe is not reliably above chance). A Not Evaluated dimension counts as non-acceptance and is shown as such.

#### Proposition 1

(False-positive contRol).

*If x*(*S*), *x*_1_, …, *x*_*m*_ *are exchangeable (in particular, under H*_0_*), then, conditional on every gating signal being available for S and for all fit runs, the probability that any gating dimension of S fails is at most r*/(*m* + 1) ≤ α .

Proof. The event that all *m* + 1 vectors are complete is symmetric in them, so conditioning on it preserves exchangeability; standardization is a symmetric function of the pooled vectors, so *M*_1_, …, *M*_*m*+1_ remain exchangeable. Some dimension fails exactly when *M*_*m*+1_ > τ, i.e. when *M*_*m*+1_ is strictly larger than at least *m* − *r* + 1 of the other *m* statistics, so that it ranks among the top *r* of the *m* + 1. By exchangeability that event has probability at most *r*/(*m* + 1). A Not Evaluated dimension is not a Fail and lies outside this bound; on honest tables it is counted separately (H3).

Three properties follow. The threshold is set by the null distribution of the same statistic on the same dataset, so no fixed cutoff is needed. Because the decision uses the maximum over all *p* signals of all three dimensions, the bound holds for the gate as a whole: adding signals cannot inflate the family-wise false-positive rate, which is the multiplicity problem a non-compensatory gate otherwise suffers. And bootstrap uncertainty enters only through the utility signals defined as interval limits; every other signal is a point estimate whose sampling variability is already represented in the null distribution. With *m* = 100 and α = 0.05, *r* = 5. The rule is taken from concurrent work [18], which compares it with compensatory alternatives; here it is used, not evaluated.

### 3.4 Confidence statements

Every gating verdict carries a statement of its own reliability, per dimension.

*Fail: strength of evidence*. For dimension *k*,

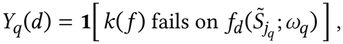

By the argument of Proposition 1, Pr(*e*_*k*_ (*S*) ≤ *u*) ≤ *u* for all *u* under exchangeability, so *e*_*k*_ is a valid conformal p-value with resolution 1/(*m* + 1). Every Fail has *e*_*k*_ ≤ *r*/(*m* + 1): if dimension *k* exceeds τ, fewer than *r* fit runs can match it.

#### Pass: limit of detection

A flaw family *f* (Table 2) is an operator that alters a fraction *d* (the *dose*) of a table’s rows in a specified way, and has a target dimension *k*(*f* ). Let π_*f*_ (*d*) be the probability that dimension *k*(*f* ) fails when *f* is applied at dose *d* to an honest table *S*_*j*_ from a null run not used for fitting. STB estimates π_*f*_ on a grid *d*_1_ > *d*_2_ > … > *d*_*L*_ from *N* independent replicates per dose, each using a distinct base run and a fresh flaw seed, and defines

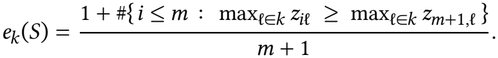

where *al* is the number of detections at *d*_*l*_ and LB_γ_ (*a, N* ) is the one-sided Clopper–Pearson lower bound at confidence γ [8]. We use π = γ = 0.95, following the clinical-laboratory convention for limits of detection [7].

**Table 2:**
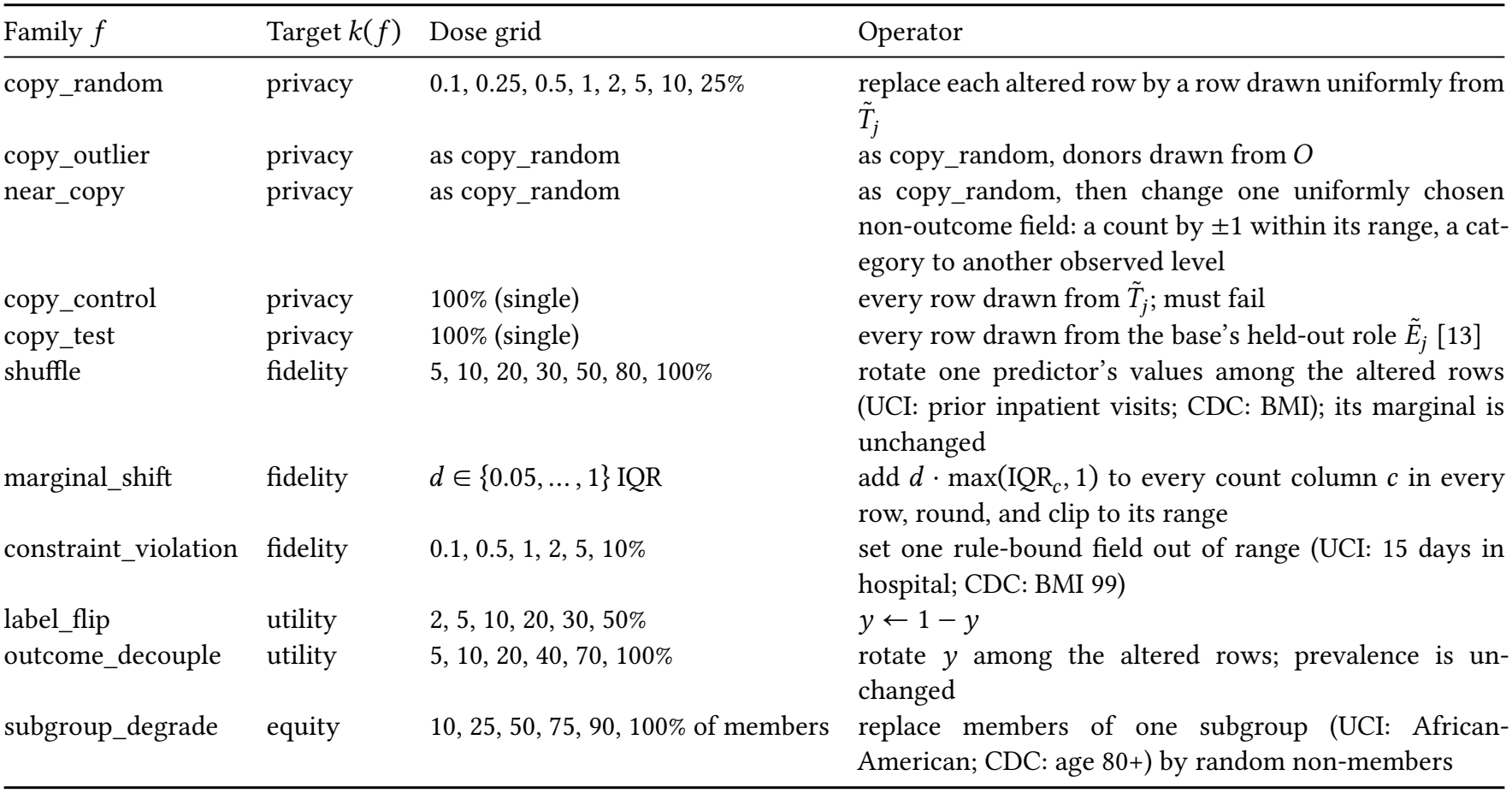
Planted flaw families. ⌈*dñ*⌉ rows are altered unless stated. 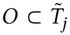 is the 5% of training rows farthest from their 10th nearest neighbour. Copy grids also include 1, 3, 10 and 30 rows wherever these are fewer rows than the smallest fractional dose. Single-dose families report a detection rate rather than a limit.

| Family *f* | Target *k*(*f*) | Dose grid | Operator |
|---|---|---|---|
| copy_random | privacy | 0.1, 0.25, 0.5, 1, 2, 5, 10, 25% | replace each altered row by a row drawn uniformly from *T̃*<sub>*j*</sub> |
| copy_outlier | privacy | as copy_random | as copy_random, donors drawn from *O* |
| near_copy | privacy | as copy_random | as copy_random, then change one uniformly chosen non-outcome field: a count by ±1 within its range, a cat-egory to another observed level |
| copy_control | privacy | 100% (single) | every row drawn from *T̃*<sub>*j*</sub>; must fail |
| copy_test | privacy | 100% (single) | every row drawn from the base's held-out role *Ẽ*<sub>*j*</sub> [13] |
| shuffle | fidelity | 5, 10, 20, 30, 50, 80, 100% | rotate one predictor's values among the altered rows (UCI: prior inpatient visits; CDC: BMI); its marginal is unchanged |
| marginal_shift | fidelity | *d* ∈ {0.05, … , 1} IQR | add *d* · max(IQR<sub>*c*</sub>, 1) to every count column *c* in every row, round, and clip to its range |
| constraint_violation | fidelity | 0.1, 0.5, 1, 2, 5, 10% | set one rule-bound field out of range (UCI: 15 days in hospital; CDC: BMI 99) |
| label_flip | utility | 2, 5, 10, 20, 30, 50% | *y* ← 1 − *y* |
| outcome_decouple | utility | 5, 10, 20, 40, 70, 100% | rotate *y* among the altered rows; prevalence is un-changed |
| subgroup_degrade | equity | 10, 25, 50, 75, 90, 100% of members | replace members of one subgroup (UCI: African-American; CDC: age 80+) by random non-members |

##### Proposition 2

(Limit of detection). *If the N replicates at each dose are independent draws of the planting procedure, then with probability at least* γ, π_*f*_ (*d*_*l*_ ) ≥ π *holds simultaneously for every l* ≤ *l*^*^.

The proof (Appendix B) uses fixed-sequence testing [40] and requires neither monotonicity of π_*f*_ in dose nor a multiplicity correction. A certificate at π = 0.95 is possible only if LB_0.95_(*N, N* ) = 0.05^1/*N*^ ≥ 0.95, i.e. *N* ≥ 59; STB uses *N* = 150 for privacy families and *N* = 100 for the others. The guarantee is conditional on the stated family, its planting procedure and the dataset; it says nothing about failure types that were not injected. It holds for each family and dataset separately, not simultaneously across them, and only at the tested grid doses; and it concerns the detector’s sensitivity to the planting procedure, not the absence of a flaw in a table that passes. A probit curve in log *d* is fitted for display only [41].

#### Validation and transfer

Proposition 2 is checked empirically: at each 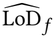, the detection rate is re-estimated on 100 fresh base runs never used for estimation. Because a synthetic table can conceal a flaw better than a real one, the privacy families are also planted into fresh generator samples that pass privacy before planting, and into the very table being judged; this table-specific limit is reported with that table’s privacy verdict, next to the honest-table limit.

#### Generators: seed agreement

For generator *g*, the verdict on each dimension is reported with the number of seeds, out of five (three for AIM on CDC; Appendix D), that agree with the modal verdict.

### 3.5 Admission of a dimension

A diagnostic dimension is promoted to gating on a dataset only if (a) its signals are available on every fit run and on every estimation base at dose 0; (b) a one-sided exact binomial test finds no excess of honest failures over α on the estimation bases; and (c) its target flaw has a certified limit of detection within the tested range. If promoted, the gate is refitted with the dimension included and every verdict, *e*_*k*_ and limit on that dataset is recomputed from the saved signals.

## 4 Experimental Design

All quantities below were fixed in a preregistration before any confirmatory table was scored; deviations are listed with dates and reasons in Appendix D. Notation follows Section 3.

### 4.1 Datasets and roles

#### CDC (primary)

The CDC Diabetes Health Indicators table (BRFSS 2015) has |*D*| = 253,680 respondents, one row each, and a binary diabetes outcome [3]. About 14% of rows exactly duplicate another respondent’s answers; they are kept, so distinct real people coincide often (*p*_0_ > 0), which tests whether a benchmark separates copying from chance agreement. The split of Section 3.1 gives |*T* | = |*P* | ≈ 101,500, |*E*| ≈ 50,700, *n* = |*R*| = |*S*| ≈ 50,700, and nullrun tables of *ñ* ≈ 40,600 rows.

#### UCI (replication)

UCI Diabetes 130-US Hospitals, earliest encounter per patient (|*D*| = 71,518), with 30-day readmission as the outcome [9, 37]: |*T* | = |*P* | ≈ 28,600, |*E*| ≈ 14,300, *n* ≈ 14,300, *ñ* ≈ 11,400.

### 4.2 Null runs

Each dataset receives 350 null runs whose indices are fixed in advance and partitioned into three disjoint sets, *J*_fit_ = {0, …, 99}, *J*_est_ = {100, …, 249} and *J*_val_ = {250, …, 349}. *J*_fit_ (*m* = 100) calibrates every verdict (Section 3.3); the honest tables 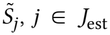, are the bases into which flaws are planted to estimate limits of detection; *j* ∈ *J*_val_ are fresh bases used only to validate them. No null run is shared with the concurrent study [18].

### 4.3 Planted flaws

A flaw family *f* maps a base table 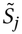, a dose *d* and a seed ω to a flawed table 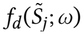 with the same columns and the same *ñ* rows. Unless stated otherwise, the dose is a fraction of rows: the seed fixes a random permutation σ of the rows, and the flaw alters rows σ_1_, …, σ_⌈*dñ* ⌉_. Because larger doses alter a superset of the rows altered by smaller ones, doses are *nested* within a replicate. Table 2 defines the families and their targets *k*(*f* ).

#### Replicates and outcome

For family *f*, replicate *q* = 1, …, *N*_*f*_ uses its own base *j*_*q*_ ∈ *J*_est_ (distinct across *q*) and its own seed ω_*q*_, and records at every grid dose

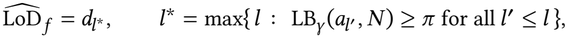

with *N*_*f*_ = 150 for privacy families and *N*_*f*_ = 100 otherwise. Dose 0 gives the false-positive rate. The detection count at *d*_*l*_ is *al* = ∑_*q*_ *Y*_*q*_ (*d*_*l*_ ), and 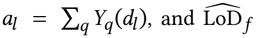 follows Section 3.4. Only *k*(*f* ) is rescored on a flawed table; the other gating dimensions are copied from the base’s dose-0 evidence. A fixed random 5% of replicates per family are re-scored in full to check empirically that this shortcut leaves the target verdict unchanged; it is not equivalent by construction, because the gate’s standardization pools all gating signals (preregistered threshold: at least 99% agreement).

#### Validation and transfer

At each 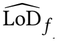 and its neighbouring grid doses, the detection rate is re-estimated on *V* = 100 fresh bases *j* ∈ *J*_val_ with fresh seeds; the claim 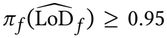 is retained if the Wilson 95% interval of the observed rate reaches 0.95. For the three graded privacy families, the same planting is repeated on fresh samples of Gaussian copula and CTGAN that pass privacy before planting; the *transfer gap* is the observed detection rate, in these tables, at the honest-table 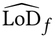.

### 4.4 Generators

Let *G* = {Gaussian copula [30], CTGAN, TVAE [42], AIM with ε = 1 [26]} and seeds *s* ∈ {17, 29, 43, 59, 71}. Each *g* ∈ *G* is fitted on *T* with default hyperparameters, and *S*_*g,s*_ of *n* rows is judged on the main roles (*T, R, E*). For every *S*_*g,s*_ that passes privacy, copy_random and copy_outlier are also planted into *S*_*g,s*_ it-self (100 replicates per dose) to give a table-specific limit. Rankings illustrate the protocol and are not a leaderboard [12]. AIM exceeded the compute budget: it was scored on CDC with seeds 29, 43 and 71 only, and not on UCI (Appendix D).

### 4.5 Comparators

Let *c*(·) be a competitor statistic from SDMetrics 0.33.0 [10], oriented so that larger is worse. It is calibrated exactly as an STB dimension with one signal: with fit values 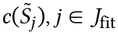, a table fails if *c*(*S*) exceeds the *r* -th largest fit value after pooled standardization. All benchmarks therefore operate under the same false-positive rule (Proposition 1). For the head-to-head, STB is restricted to the target dimension alone, STB_*k*(*f* )_, so that neither side gains from the other dimensions’ signals.

- **Primary comparators (whole table)**. *c*_priv_ = 1 − New-RowSynthesis, the share of synthetic rows that match a training row (NewRowSynthesis computed by a vectorized implementation of SDMetrics’ exact rule, verified to agree exactly); *c*_fid_ = 1 − the Quality Report score. Both are scored on copy_random, near_copy, shuffle and marginal_shift, on the first 100 replicates of each, and every benchmark’s limit in this comparison, STB’s included, is computed from exactly those replicates.
- **DCR comparison (supporting, detection rates only)**.

DCRBaselineProtection and DCROverfittingProtection are scored at *d* ∈ {0, 1%, 5%, 25%} of copy_random and near_copy on 30 replicates. Their built-in subsampling compares *b* = 2,000 released rows with *b* training rows, so STB’s privacy dimension is also scored at budget *b* (STB^(*b*)^), calibrated on *b*-row samples of the fit runs.

SynthEval was not included because its configuration could not be fixed before the preregistration.

### 4.6 Hypotheses

**H1** On each dataset, at the same false-positive rule, STB’s privacy dimension calibrated on its own is non-inferior to NewRowSynthesis on copy_random and superior on near_copy:

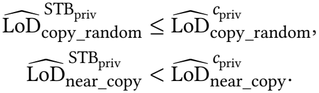

Limits are grid doses estimated from the same 100 replicates; a family without a certified limit counts as above the largest dose, and two uncertified limits tie. Predicted before scoring: a tie on copy_random, because NewRow-Synthesis counts exact matches; STB lower on near_copy, because one changed field defeats exact matching; competitor limits higher on CDC, where *p*_0_ is large; and DCR blind until a large share of rows is copied, because it compares median distances. No advantage is predicted for fidelity.

**H2** (descriptive) On fully re-scored replicates, the rate at which each gating dimension fails, by family and dose; co-firing of a non-target dimension is reported, not counted as an error.

**H3** (design check) On each dataset, the number of honest tables 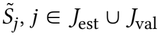, *j* ∈ *J*_est_ ∪ *J*_val_ (250 tables), on which the gate fails is tested against the bound *r*/(*m* + 1) = 5/101 with an exact one-sided binomial test at level 0.05; H3 holds if neither dataset’s test rejects. Each gating dimension’s Fail rate is reported with an exact one-sided 95% upper bound, and Not Evaluated verdicts are counted separately. A gate refitted after admitting equity is tested on *J*_val_ only, because *J*_est_ decided its admission.

**H4** (risky) For each benchmark and dimension, generators are ranked within each seed by the dimension’s statistic (max_*l*∈*k*_ *z*_*l*_ for STB), and Kendall’s rank correlation τ_*b*_ is computed between the rankings of every pair of seeds. A ranking is *unstable* if the mean τ_*b*_ is below 2/3, i.e. if on average more than one in six generator pairs reverses order between two seeds. H4 predicts instability under every benchmark.

We also predicted that copy_test is missed by every benchmark, STB included: a release of real held-out records has no excess matches with *T* and real-data fidelity. H3 concerns the false-positive rate conditional on the 100 fit runs, which all 250 honest tables share. That conditional rate varies with the fit runs. For scores drawn from a fixed continuous distribution it would follow Beta(*r, m* − *r* + 1) = Beta(5, 96), with mean 5/101, and an exactly calibrated gate would then fail the H3 test on a given dataset with probability about 0.16; pooled standardization makes this an approximation (computed before scoring results; not preregistered).

## 5 Results

### 5.1 Calibration holds on honest tables (H3)

On the 250 honest estimation and validation tables, the gate failed 7 tables on CDC (2.8%; exact one-sided binomial test against 5/101, *p* = 0.97) and 8 on UCI (3.2%, *p* = 0.93), so H3 holds on both datasets. Fidelity failed 3 and 7 tables (exact one-sided 95% upper bounds 3.1% and 5.2%), utility 4 and 1, and privacy none on either dataset (upper bound 1.2%). No honest table was Not Evaluated. The test detects excess failures rather than certifying the bound, but both observed rates lie below 4.95%. These tables have the null-run size; no honest table was scored at the 1.25-times-larger generator size, so for generator verdicts the bound remains an approximation.

### 5.2 Limits of detection

Table 3 lists the certified limits. Privacy limits differ sharply between the datasets, in the direction set by chance agreement. On UCI, where distinct patients almost never share a row, STB certified copy_random and copy_outlier at 3 copied rows out of 11,443 (0.026%); a single copied row was caught in 15% of replicates. On CDC, where about 14% of respondents duplicate another respondent exactly, copies hide among chance matches and both families were certified at 2% (812 of 40,589 rows). near_copy, which alters one field of each copied row, was certified at 10% on UCI and not detected reliably at any tested dose on CDC. Among the fidelity and utility families, constraint_violation and label_flip were detected at the lowest tested dose on both datasets, so their limits are upper bounds; shuffle, which preserves every marginal, was certified at 30% on CDC and not detected on UCI; outcome_decouple needed 10% on CDC and 70% on UCI. copy_control was detected in all 150 replicates on both datasets and copy_test in none, as predicted.

**Table 3:**
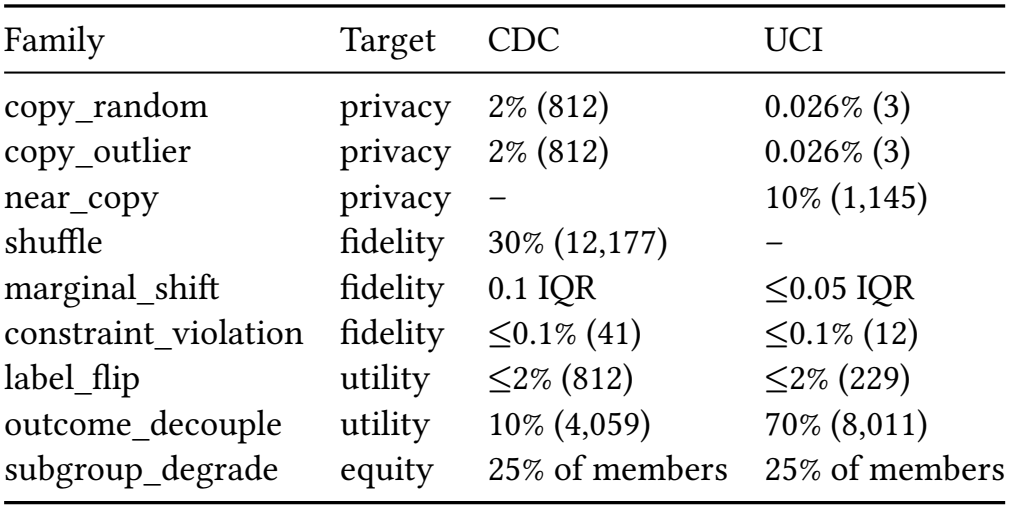
Certified limits of detection on honest tables: fraction of rows (rows). *n* = 40,589 (CDC), 11,443 (UCI). ≤: detected at the lowest tested dose. –: not detected with probability ≥ 0.95 at any tested dose.

### 5.3 Do the stated limits hold?

On the 100 fresh validation bases, every certified family on both datasets was detected in at least 99 of 100 replicates at its limit (100/100 in 15 of 16 cases). The one-sided 95% Clopper–Pearson lower bound was at least 0.953 in every case, so each limit was certified again on independent data, beyond the preregistered Wilson compatibility check. The limits did not transfer unchanged to synthetic tables. On CDC, copies planted at the honest-table limit of 2% into Gaussian copula and CTGAN tables that passed privacy were detected in at most 1 of 150 replicates; in those tables the limit was 10%. The table-specific limits agree: copies planted into the very table being judged were certified at 5–10% of its rows for Gaussian copula, CTGAN and TVAE, and at 1–5% for AIM. On UCI the 3-row limit held in every Gaussian copula and CTGAN table, and near_copy at the honest-table limit was detected in 98% (CTGAN) and 87% (Gaussian copula) of replicates.

### 5.4 STB against SDMetrics (H1)

Table 4 compares limits on the same 100 replicates under the same calibration rule. On UCI, STB’s privacy dimension tied NewRow-Synthesis on copy_random (3 rows each) and certified near_copy at 5%, which NewRowSynthesis never detected, so H1 holds. New-RowSynthesis measures exact row novelty by definition, so its blindness to near_copy is expected; the result supports combining exact-match and proximity signals, not superiority over privacy-evaluation frameworks in general. On CDC it does not: NewRowSynthesis certified copy_random at 0.5% against STB’s 1%, and neither detected near_copy. The same rule did not give the same realized false-positive rate on these bases: NewRowSynthesis failed 6 of 100 unflawed tables on CDC and none on UCI, STB’s privacy dimension 2 and 9. Contrary to our prediction, STB was more sensitive on fidelity. The Quality Report needed a shift of 0.2 IQR on both datasets and never detected shuffle, against 0.1 and ≤0.05 IQR and 30% (CDC) for STB. DCRBaselineProtection and DCROverfitting-Protection detected 0–13% of replicates at every dose, including 25% copying, whereas STB as shipped detected 1% copying in all 30 replicates on both datasets. At DCR’s 2,000-row budget, STB detected 1% copying in 27 of 30 replicates on UCI but needed 25% on CDC (29 of 30).

**Table 4:** Head-to-head limits on the same 100 replicates under the same calibration rule. STB: the target dimension alone. NRS: NewRowSynthesis; QR: Quality Report. –: not detected.

|  | CDC |  | UCI |  |
|---|---|---|---|---|
| Family | STB | SDMetrics | STB | SDMetrics |
| copy_random | 1% | 0.5% (NRS) | 3 rows | 3 rows (NRS) |
| near_copy | – | − (NRS) | 5% | − (NRS) |
| marginal_shift | 0.1 IQR | 0.2 IQR (QR) | ≤0.05 IQR | 0.2 IQR (QR) |
| shuffle | 30% | − (QR) | – | − (QR) |

### 5.5 Real generators (exploratory)

Generator tables are 1.25 times larger than the calibration tables, so Proposition 1 does not formally cover them, and we report these verdicts as an exploratory application of the protocol. Every fidelity and utility Fail lies at or near the largest standardized value the calibration allows, far from the threshold; this suggests, but does not establish, that these verdicts would survive calibration at the matching size.

Every generator failed fidelity and utility on every seed of both datasets, each with the smallest possible evidence strength, *e*_*k*_ = 1/101: each table was more extreme than all 100 honest calibration tables (Table 5). Privacy separated them. On CDC every table passed privacy. On UCI, Gaussian copula and CTGAN passed on all five seeds, whereas TVAE failed on all five, again with *e* = 1/101. The Fail came from proximity, not copying: TVAE’s proximity share exceeded its real-data floor by 17–22 percentage points, against 1–2 for CTGAN and 0 for Gaussian copula, while exact copies were absent on three seeds and 0.014% of rows on the other two. On the same tables the distance-based membership attack scored an AUC of 0.497–0.505, at chance. The indicators disagree, and the Fail is a proximity anomaly, not demonstrated leakage: TVAE also fails fidelity, and concentrated samples can lie close to training patients without giving an attacker membership advantage. Whether the proximity is specific to training patients, rather than to real patients in general, is the test that would decide it.

**Table 5:**
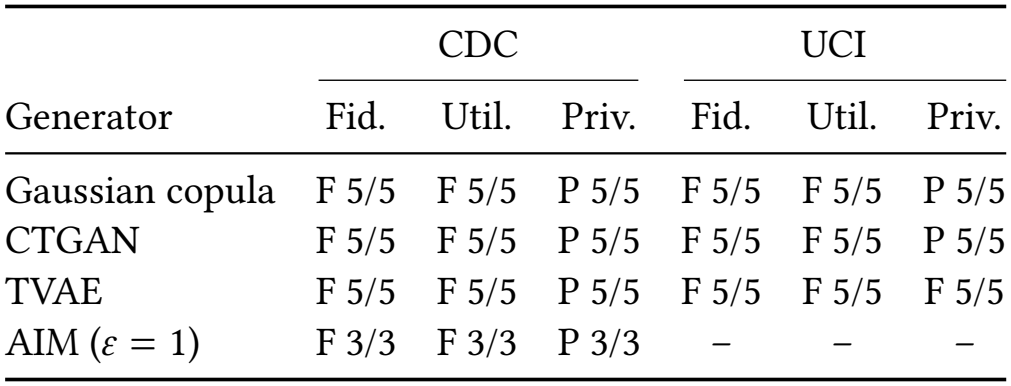
Verdicts on real generator tables, with the number of seeds agreeing. F: Fail (*e*_*k*_ = 1/101 in every case); P: Pass. AIM: three seeds, CDC only.

Every gating verdict agreed across all seeds (5/5, and 3/3 for AIM). Rankings did not. Generators were ranked within each seed and compared across seed pairs by Kendall’s τ_*b*_ over the generators both seeds contain. Under STB, privacy and utility rankings were unstable on CDC (mean τ_*b*_ = 0.13 and 0.07), and utility on UCI (0.47); fidelity rankings were stable (0.93 and 1.00). Under SDMetrics, the Quality Report ranking was stable (1.00 and 0.73); DCR and NewRowSynthesis rankings were unstable on CDC (0.52–0.60), and on UCI NewRowSynthesis produced ties that left a single usable seed pair. H4, which predicted instability under every benchmark, therefore holds only in part. The stable fidelity rankings carry little information: every table failed fidelity at or near the largest standardized value the pooled calibration allows, so their order reflects differences far beyond the threshold.

### 5.6 What else fires (H2) and equity admission

On fully re-scored replicates, flaws often failed a second dimension. On CDC, copy_outlier also failed fidelity from 1–2% of rows; near_copy failed fidelity, not privacy, at 10–25%; and constraint_violation and marginal_shift also failed utility. The target-only shortcut agreed with full re-scoring on at least 99% of replicates for every family except shuffle on UCI (39 of 40, below the preregistered 99%); the UCI shuffle result, a non-detection, therefore describes the targetonly procedure, not the full gate. Equity was not admitted on either dataset. It failed criterion (a), because its subgroup signals were not available on every fit run and estimation base, although it showed no excess of honest failures and had a certified limit (25% of a subgroup’s members).

## 6 Discussion

### What a pass means

An STB Pass is a measured statement about the benchmark, not a guarantee about the table: under the specified planting procedure, a flaw at each certified grid dose, from the largest tested dose down to the stated limit, is detected with probability at least 0.95. Without monotonicity, nothing is certified between grid doses, and sensitivity to planted flaws is not a confidence statement about how much contamination a passing table contains. The limit that matters is the one measured in the kind of table being judged. In our exploratory generator application, on UCI, a Gaussian copula or CTGAN table that passes privacy carries a table-specific limit of 3 copied rows; on CDC, the table-specific limits are 5–10% of rows, five times the honest-table limit. A Pass therefore says how large a copying failure of the planted kind would have to be to be caught reliably, not that none is present.

### Where STB ties or loses

On CDC, NewRowSynthesis certified copy_random at half of STB’s dose, and neither caught near_copy; no benchmark caught copy_test, a release of real held-out records; and limits measured in honest tables were optimistic for generator output on CDC, by a factor of five. Reporting these cases is part of the claim: a limit of detection is useful precisely because it says where a check stops working.

### Limits of detection as a reporting standard

Any benchmark that issues pass/fail verdicts can be calibrated and characterized this way: the null runs and planted flaws need only the evaluator’s own statistics. Clinical laboratories made the limit of detection a precondition for using an assay [7]; we argue synthetic-data benchmarks should do the same for every verdict.

### Relation to the critiques

STB does not dispute that similaritybased privacy metrics and fidelity scores can mislead [13, 33]; it measures how much, for each check, and reports the answer with the verdict.

## 7 Limitations

STB covers structured tabular data and binary prediction on two datasets. Its limits of detection hold only for the flaw families we planted, planted as specified; real failures may look different, which is why we report the transfer gap and check disagreements on real generator output directly. The membership attack is distance-based and gives a lower bound on risk, not a guarantee. The false-positive bound is marginal over the choice of fit runs; conditional on one set of fit runs the rate varies around 5/101. A Fail means a departure from real data detectable at the tested size, not clinical harm: that every generator failed fidelity and utility shows separation from the real-data null, not that the tables are unusable for a given task. Equity and robustness do not vote; equity was not admitted on either dataset, so the gate covers three of the five dimensions. Calibration uses pseudo-roles drawn from one cohort per dataset, and real generator tables are 1.25 times larger than the null tables. Computational constraints prevented calibration at the generator tables’ size; the formal false-positive guarantee therefore applies only to the matched-size protocol, and generator results are exploratory. Planted flaws re-score only their target dimension, a shortcut checked empirically rather than exact. Generators use default hyperparameters, and AIM was evaluated on three seeds on CDC only, seeds whose fits finished within the compute budget. We compare against one external benchmark. The audit is small and was coded by one author.

## 8 Conclusion

STB treats every benchmark verdict as a measurement that must be calibrated and characterized before it is trusted: its false-positive rate is bounded on real tables of the calibration size, and each Pass states the smallest planted flaw it would have caught. On two diabetes datasets, honest tables failed at 2.8% and 3.2%, within the bound; STB certified privacy limits as low as 3 copied rows, and, in an exploratory application, it flagged a proximity anomaly in TVAE tables that a membership-inference attack did not register. We release the code, null-run identifiers and preregistration so that other benchmarks can be held to the same standard.

### A Reproducibility

Code, the preregistration, the saved evidence of every scored table, and scripts that recompute every number and figure in this paper are available at https://anonymous.4open.science/r/SynTrustBench_v05/. Every evaluated table stores its raw signals, so verdicts, evidence and limits are functions of saved evidence. Each dimension draws from its own random stream, spawned from a per-table seed, so scoring one dimension alone reproduces its value inside a full profile. A manifest records input hashes, the plan, code hashes and package versions, and refuses to mix platforms or code versions; all confirmatory runs were scored on one Linux x86-64 server.

### B Proof of Proposition 2

Order the doses *d*_1_ > *d*_2_ > … > *d*_*L*_ and let *H*_*l*_ be the hypothesis that the detection probability at *d*_*l*_ is below π . The procedure rejects *H*_1_, *H*_2_, … in order while the one-sided Clopper–Pearson lower bound at level γ is at least π, and stops at the first non-rejection; the rejected hypotheses therefore form a prefix. Suppose some reported claim is false, and let *l*_0_ be the first index in the order at which 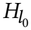 is true. Because rejections form a prefix and a false claim was reported at or after *l*_0_, 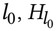 was rejected. The Clopper–Pearson test of a single true 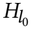 has size at most 1 − γ when its replicates are independent draws. Hence *P* (some reported claim is false) ≤ *P* (reject 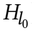) ≤ 1 − γ . No monotonicity in dose and no multiplicity correction were used.

### C Signal definitions

#### Probes

Random forest: 200 trees, minimum leaf size 5. Logistic regression: *C* = 1, up to 3,000 iterations. Gradient boosting: histogram gradient boosting with 200 iterations and native categorical handling. Random forest and logistic regression use one-hot categories and standardized continuous columns; all encoders are fitted on *T* only, and no hyperparameter is tuned, so no information from *E* enters training. **Two-sample classifier**. Histogram gradient boosting (200 iterations), 5-fold stratified cross-fitting; AUC on out-of-fold scores. **Marginal distances**. *W*_1_ between empirical distributions, divided by the interquartile range of the column in *R*; Jensen–Shannon divergence in bits between category frequencies. **Intervals**. Percentile bootstrap, *B* = 200, 95%; utility intervals resample rows of *E* jointly for the synthetic-and real-trained probes (paired). **Privacy**. Nearest neighbours by exact Euclidean search within reproducible samples of at most 10,000 rows of *T*, of *S* and of *E*; exact matches over all rows after canonical formatting. **Equity**. Subgroups with at least 30 test patients and at least 10 of each outcome; retention as in Section 3.2, against the subgroup’s real-trained AUC. **Calibration**. *m* = 100 fit runs, α = 0.05, *r* = 5, scale floor η = 10^−8^. Full code: stb/ in the artifact (Appendix A).

### D Preregistration and deviations

The preregistration (PREREGISTRATION_v0.5.md in the artifact) was frozen on 12 November 2026, before any confirmatory table was scored. Its SHA-256 hash is

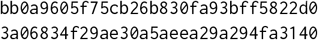

Before freezing, its hypotheses were revised on 11 November so that each is testable as stated: H1 became non-inferiority on copy_random and superiority on near_copy, H3 an exact binomial test against 5/101, and H4 a fixed threshold τ_*b*_ < 2/3. The revision log is part of the file. Deviations after freezing:

1. *AIM on CDC* (15 November). Fits for seeds 17 and 59 were stopped after more than 60 hours; AIM is reported on seeds 29, 43 and 71. Because the surviving seeds are those whose fits finished, they may not be representative. For H4, Kendall’s τ_*b*_ is computed for each seed pair over the generators both seeds contain: four for pairs of AIM’s seeds, three other-wise.
2. *AIM on UCI* (15 November). Because AIM fits on CDC took from 11 to more than 60 hours, AIM was not started on UCI; UCI results cover three generators.

The stopped and skipped AIM fits are being rerun with the pre-registered settings; their results will be added to the final version, together with H4 recomputed on all available seeds. The number of parallel workers was changed during the runs; each table draws from its own seeded random streams, so results do not depend on it.

## Data Availability

This study used two publicly available, de-identified datasets: CDC Diabetes Health Indicators (BRFSS 2015), https://archive.ics.uci.edu/dataset/891/cdc+diabetes+health+indicators, and Diabetes 130-US Hospitals for Years 1999-2008 (doi:10.24432/C5230J), https://archive.ics.uci.edu/dataset/296/diabetes+130-us+hospitals+for+years+1999-2008. All data produced in the present study, including the SynTrustBench v0.5 code, the frozen preregistration, null-run identifiers, the saved evidence for every scored table, and scripts that regenerate every number and figure in the manuscript, are available online at https://anonymous.4open.science/r/SynTrustBench_v05/

## Generative AI Disclosure

The authors used Anthropic’s Claude to edit and restructure prose, formalize notation and proofs, draft and review analysis and experiment-orchestration code, and check consistency between the paper, the preregistration and the code. The study design, hypotheses, preregistration and interpretation are the authors’. All code was run, and all numbers were produced, by the released pipeline; the authors checked every generated text and take full responsibility for the content.

## References

[1] Brian Belgodere, Pierre Dognin, Adam Ivankay, Igor Melnyk, Youssef Mroueh, Aleksandra Mojsilović, Jiri Navratil, Apoorva Nitsure, Inkit Padhi, Mattia Rigotti, Jerret Ross, Yair Schiff, Radhika Vedpathak, and Richard A. Young. 2024. Auditing and Generating Synthetic Data With Controllable Trust Trade-Offs. IEEE Journal on Emerging and Selected Topics in Circuits and Systems 14, 4 (2024), 773–788. doi:10.1109/JETCAS.2024.3477976

[2] Mohammadreza Beyki. 2021. SCorGAN: Generating Synthetic Healthcare Records with Convolutional Generative Adversarial Networks. Master’s thesis. Virginia Polytechnic Institute and State University.

[3] Centers for Disease Control and Prevention. 2015. CDC Diabetes Health Indicators. UCI Machine Learning Repository, dataset 891.

[4] Taha Ceritli, Ghadeer O. Ghosheh, Vinod Kumar Chauhan, Tingting Zhu, Andrew P. Creagh, and David A. Clifton. 2023. Synthesizing Mixed-type Electronic Health Records using Diffusion Models. arXiv:2302.14679 [cs.LG] doi:10.48550/arXiv.2302.14679

[5] Richard J. Chen, Ming Y. Lu, Tiffany Y. Chen, Drew F. K. Williamson, and Faisal Mahmood. 2021. Synthetic data in machine learning for medicine and healthcare. Nature Biomedical Engineering 5 (2021), 493–497. doi:10.1038/s41551-021-00751-8

[6] Xingran Chen, Zhenke Wu, Xu Shi, Hyunghoon Cho, and Bhramar Mukherjee. 2025. Generating Synthetic Electronic Health Record Data: A Methodological Scoping Review with Benchmarking on Phenotype Data and Open-Source Software. Journal of the American Medical Informatics Association (2025). doi:10.1093/jamia/ocaf082

[7] Clinical and Laboratory Standards Institute. 2012. Evaluation of Detection Capability for Clinical Laboratory Measurement Procedures; Approved Guideline— Second Edition. CLSI.

[8] C. J. Clopper and E. S. Pearson. 1934. The Use of Confidence or Fiducial Limits Illustrated in the Case of the Binomial. Biometrika 26, 4 (1934), 404–413.

[9] John Clore, Krzysztof J. Cios, Jonathan P. DeShazo, and Beata Strack. 2014. Diabetes 130-US Hospitals for Years 1999–2008. UCI Machine Learning Repository. doi:10.24432/C5230J

[10] DataCebo. [n. d.]. SDMetrics: Metrics for Synthetic Data Generation Projects. https://github.com/sdv-dev/SDMetrics. Version 0.33.0 used.

[11] Damien Desfontaines. [n. d.]. Better Empirical Privacy Metrics. https://desfontain.es/blog/better-empirical-privacy-metrics.html.

[12] Yuntao Du and Ninghui Li. 2024. Towards Principled Assessment of Tabular Data Synthesis Algorithms. SynMeter. arXiv:2402.06806

[13] Georgi Ganev and Emiliano De Cristofaro. 2025. The Inadequacy of Similarity-based Privacy Metrics: Privacy Attacks against “Truly Anonymous” Synthetic Datasets. In IEEE Symposium on Security and Privacy (SP). arXiv:2312.05114

[14] Georgi Ganev and Emiliano De Cristofaro. 2025. Synthetic Data, Similarity-based Privacy Metrics, and Regulatory (Non-)Compliance. arXiv:2505.01524

[15] Yijie Hao, Huan He, and Joyce C. Ho. 2024. LLMSYN: Generating Synthetic Electronic Health Records Without Patient-Level Data. In Proceedings of the 9th Machine Learning for Healthcare Conference (Proceedings of Machine Learning Research, Vol. 252).

[16] Frederik Harder, Kamil Adamczewski, and Mijung Park. 2021. Differentially Private Mean Embeddings with Random Features for Generative Modeling. In Proceedings of the 24th International Conference on Artificial Intelligence and Statistics (Proceedings of Machine Learning Research, Vol. 130). 1819–1827.

[17] Neeam Shahriar Hayder and Syed Ahmad Chan Bukhari. 2026. SynTrustBench: An Evidence-Gated and Executable Benchmark for Trustworthiness Claims in Synthetic Clinical Data. medRxiv preprint. doi:10.64898/2026.08.05.26359803

[18] Neeam Shahriar Hayder, Behraj Khan, and Syed Ahmad Chan Bukhari. 2026. A Calibrated Gate for Evaluating Synthetic Clinical Data. Preprint.

[19] Mikel Hernandez, Gorka Epelde, Ane Alberdi, Rodrigo Cilla, and Debbie Rankin. 2022. Synthetic data generation for tabular health records: A systematic review. Neurocomputing 493 (2022), 28–45. doi:10.1016/j.neucom.2022.04.053

[20] Bing Hu, Yixin Li, Asma Bahamyirou, and Helen Chen. 2025. SynQP: A Framework and Metrics for Evaluating the Quality and Privacy Risk of Synthetic Data. In 2025 22nd Annual International Conference on Privacy, Security, and Trust (PST). 1–6. doi:10.1109/PST65910.2025.11268831

[21] James Jordon, Jinsung Yoon, and Mihaela van der Schaar. 2019. PATE-GAN: Generating Synthetic Data with Differential Privacy Guarantees. In International Conference on Learning Representations.

[22] Bayrem Kaabachi, Jérémie Despraz, Thierry Meurers, Karen Otte, Mehmed Halilovic, Bogdan Kulynych, Fabian Prasser, and Jean Louis Raisaro. 2025. A scoping review of privacy and utility metrics in medical synthetic data. npj Digital Medicine 8 (2025), 60. doi:10.1038/s41746-024-01359-3

[23] Francis John Kita, Gadde Srinivasa Rao, and Peter Josephat Kirigiti. 2025. A deep learning framework for predicting cardiovascular multimorbidity using synthetic data. Connection Science 37, 1 (2025), 2565163. doi:10.1080/09540091.2025.2565163

[24] Anantaa Kotal and Anupam Joshi. 2024. ContextGAN: Mitigating Contextual Privacy Leakage with Differential Privacy for Synthetic Data. In 2024 IEEE International Conference on Big Data. 6289–6297. doi:10.1109/BigData62323.2024.10826047

[25] Anton D. Lautrup, Tobias Hyrup, Arthur Zimek, and Peter Schneider-Kamp. 2024. SynthEval: A Framework for Detailed Utility and Privacy Evaluation of Tabular Synthetic Data. Data Mining and Knowledge Discovery (2024).

[26] Ryan McKenna, Brett Mullins, Daniel Sheldon, and Gerome Miklau. 2022. AIM: An Adaptive and Iterative Mechanism for Differentially Private Synthetic Data. Proceedings of the VLDB Endowment 15, 11 (2022), 2599–2612.

[27] Ivana Nanevski, Maryam Mohebi, Sebastian Jäger, Karen Otte, Fabian Prasser, Matthias Schulte-Althoff, Daniel Fürstenau, and Felix Biessmann. 2026. Evaluating the quality of tabular synthetic data in health care. PLOS Digital Health 5, et al. 7 (2026), e0001522. doi:10.1371/journal.pdig.0001522

[28] Ahmed Ammar Naseer, Benjamin Walker, Christopher Landon, Andrew Ambrosy, Marat Fudim, Nicholas Wysham, Botros Toro, Sumanth Swaminathan, and Terry Lyons. 2023. ScoEHR: Generating Synthetic Electronic Health Records using Continuous-time Diffusion Models. In Proceedings of the 8th Machine Learning for Healthcare Conference (Proceedings of Machine Learning Research, Vol. 219). 489–508.

[29] National Institute of Standards and Technology. [n. d.]. SDNist: Synthetic Data Report Tool and the CRC Data and Metrics Archive. https://github.com/usnistgov/SDNist.

[30] Neha Patki, Roy Wedge, and Kalyan Veeramachaneni. 2016. The Synthetic Data Vault. In 2016 IEEE International Conference on Data Science and Advanced Analytics. 399–410. doi:10.1109/DSAA.2016.49

[31] Yue Qi, Lorrie Herbault, Hadrien Lautraite, Michael Yu, Katleen Blanchet, Christian Vincelette, Louis Mullie, Guillaume Dumas, Jean-François Rajotte, Kamran Afzali, Sébastien Gambs, and Michaël Chassé. 2026. CLOVER: A framework for benchmarking synthetic data generation methods balancing utility and privacy in healthcare. Artificial Intelligence in the Life Sciences 9 (2026), 100155. doi:10.1016/j.ailsci.2026.100155

[32] Zhaozhi Qian, Rob Davis, and Mihaela van der Schaar. 2023. Synthcity: A benchmark framework for diverse use cases of tabular synthetic data. In Advances in Neural Information Processing Systems, Vol. 36. 3173–3188. doi:10.52202/075280-0140

[33] Ossi Räisä, Boris van Breugel, and Mihaela van der Schaar. 2025. Position: All Current Generative Fidelity and Diversity Metrics are Flawed. In Proceedings of the 42nd International Conference on Machine Learning (Proceedings of Machine Learning Research, Vol. 267). 82016–82050.

[34] Resmi Ramachandranpillai, Md Fahim Sikder, David Bergström, and Fredrik Heintz. 2024. BT-GAN: Generating Fair Synthetic Healthdata via Bias-transforming Generative Adversarial Networks. Journal of Artificial Intelligence Research 79 (2024), 1313–1341. doi:10.1613/jair.1.15317

[35] Theresa Stadler, Bristena Oprisanu, and Carmela Troncoso. 2022. Synthetic Data – Anonymisation Groundhog Day. In 31st USENIX Security Symposium (USENIX Security 22). 1451–1468.

[36] Thomas Steinke, Milad Nasr, and Matthew Jagielski. 2023. Privacy Auditing with One (1) Training Run. In Advances in Neural Information Processing Systems (NeurIPS), Vol. 36.

[37] Beata Strack, Jonathan P. DeShazo, Chris Gennings, Juan L. Olmo, Sebastian Ventura, Krzysztof J. Cios, and John N. Clore. 2014. Impact of HbA1c Measurement on Hospital Readmission Rates: Analysis of 70,000 Clinical Database Patient Records. BioMed Research International 2014 (2014), 781670. doi:10.1155/2014/781670

[38] Amirsina Torfi and Edward A. Fox. 2020. CorGAN: Correlation-Capturing Convolutional Generative Adversarial Networks for Generating Synthetic Healthcare Records. In Proceedings of the Thirty-Third International Florida Artificial Intelligence Research Society Conference. 335–340. doi:10.32473/flairs.v33i.128262

[39] Vibeke Binz Vallevik, Aleksandar Babić, Serena Elizabeth Marshall, Severin Elvatun, Helga Brøgger, Sharmini Alagaratnam, Bjørn Edwin, Narasimha Raghavan Veeraragavan, Anne Kjersti Befring, and Jan Franz Nygård. 2024. Can I trust my fake data—A comprehensive quality assessment framework for synthetic tabular data in healthcare. International Journal of Medical Informatics 185 (2024), 105413. doi:10.1016/j.ijmedinf.2024.105413

[40] Peter H. Westfall and Alok Krishen. 2001. Optimally Weighted, Fixed Sequence and Gatekeeper Multiple Testing Procedures. Journal of Statistical Planning and Inference 99, 1 (2001), 25–40.

[41] Westgard QC. [n. d.]. Probit Analysis for Detection Limits. https://westgard.com/probit-part-one.htm. Secondary explainer for the probit LoD approach.

[42] Lei Xu, Maria Skoularidou, Alfredo Cuesta-Infante, and Kalyan Veeramachaneni. 2019. Modeling Tabular Data using Conditional GAN. In Advances in Neural Information Processing Systems, Vol. 32.

[43] Chao Yan, Yao Yan, Zhiyu Wan, Ziqi Zhang, Larsson Omberg, Justin Guinney, Sean D. Mooney, and Bradley A. Malin. 2022. A multifaceted benchmarking of synthetic electronic health record generation models. Nature Communications 13 (2022), 7609. doi:10.1038/s41467-022-35295-1

[44] Ghada Zamzmi, Adarsh Subbaswamy, Elena Sizikova, Edward Margerrison, Jana G. Delfino, and Aldo Badano. 2025. Scorecard for synthetic medical data evaluation. Communications Engineering 4 (2025), 130. doi:10.1038/s44172-025-00450-1

